# Efficacy of clarithromycin on COVID-19 pneumonia without oxygen administration; protocol for multicenter, open-label, randomized-controlled, 3-armed parallel group comparison, exploratory trial (CAME COVID study)

**DOI:** 10.1101/2021.06.02.21258257

**Authors:** Kazuko Yamamoto, Naoki Hosogaya, Noriho Sakamoto, Haruo Yoshida, Hiroshi Ishii, Kazuhiro Yatera, Koichi Izumikawa, Katsunori Yanagihara, Hiroshi Mukae

## Abstract

**Introduction:** The coronavirus disease 2019 (COVID-19) epidemic has been emerged worldwide. Although several medications have been approved for treating moderate-to-severe COVID-19, no treatment strategy has been established for mild COVID-19 patients who do not require oxygen administration. The spread of SARS -CoV-2 has been mostly through patients with mild COVID-19; therefore, treating patients with mild COVID-19 is critical in society. Clarithromycin is a macrolide antimicrobial agent that has been widely used for bacterial respiratory infectious diseases. Clarithromycin also acts an immunomodulating drug and suppresses cytokine storms in viral respiratory diseases, including influenza infection. In this study, we aimed to evaluate the efficacy of clarithromycin in patients with mild COVID-19.

**Methods and analysis:** This is a multicenter, open-label, randomized controlled, 3-armed parallel group comparison, exploratory trial. Subjects with mild COVID-19 pneumonia who did not require oxygen administration were enrolled and randomly assigned in a 1:1:1 ratio to Group A (administration of clarithromycin 800 mg/day), Group B (administration of clarithromycin 400 mg/day), or Group C (standard treatment without clarithromycin). The primary endpoint was the number of days required to improve clinical symptoms as measured by the severity score. Secondary endpoints included days to recover the body temperature, proportion of subjects with oxygen administration, inflammatory cytokines, viral load, serum immunoglobulins, peripheral blood lymphocytes, blood biomarkers, and pneumonia infiltrations.

**Ethics and dissemination:** The study protocol was approved by the Clinical Research Review Board of Nagasaki University in accordance with the Clinical Trials Act in Japan. The study will be conducted in accordance with the Declaration of Helsinki, the Clinical Trials Act, and other current legal regulations in Japan. Written informed consent will be obtained from all participants. The results of this study will be reported as journal publications.

**Registration:** This study was registered at the Japan Registry of Clinical Trials (registration number: jRCTs071210011).

**Strengths and limitations of this study:**

- This is the first randomized controlled trial to evaluate the efficacy of clarithromycin against COVID-19 pneumonia, especially in patients with mild COVID-19 pneumonia who do not require oxygen administration.
- To date, no treatment strategy has been established for mild COVID-19 pneumonia.
- The major limitations of this study are its exploratory nature and relatively small sample size.
- Another limitation is the open-label study design and generalizability because this study was conducted only in Japan with Japanese patients.

## Introduction

The coronavirus disease 2019 (COVID-19) epidemic is currently a major concern worldwide. In Japan, a cumulative of 562,462 polymerase chain reaction (PCR) test-positive cases have been confirmed and 9,910 deaths were reported by the Ministry of Health, Labour and Welfare in Japan as of April 26th, 2021^1^. Approximately 20% of patients with COVID-19 were hospitalized, 5% had severe symptoms requiring intensive care, and 2% were fatal in Japan^2^. Recently, dexamethasone and remdesivir have been used as standard treatments for moderate-to-severe COVID-19 patients who require respiratory support^3-5^. However, no medical treatment has been established for mild COVID-19 disease, which accounts for the majority (approximately 80%) of COVID-19 patients, and this may increase the risk of spreading severe acute respiratory syndrome coronavirus 2 (SARS-CoV-2). Although the SARS-CoV-2 vaccination program has been introduced worldwide, the concern of variant viral spread could be another challenge. Moreover, there is no treatment available to prevent exacerbations in patients with mild COVID-19^3^, which may be a critical concern during the spread of SARS-CoV-2.

The mechanism of exacerbation in coronavirus disease has been reported to correlate with dysregulation of the immune response, resulting in exaggerated inflammation to produce excessive cytokines (the so-called cytokine storm) ^6^. Indeed, infection with SARS-CoV-2 induces high expression of inflammation-related cytokines, such as granulocyte macrophage colony-stimulating factor and interleukin-6 (IL-6), accelerating inflammation^7^. Therefore, suppression of inflammatory cytokines is an important target for preventing the exacerbation of COVID-19.

Clarithromycin is a macrolide antibiotic that has been widely used as a monotherapy for bacterial respiratory infectious diseases. Clarithromycin has also been used as a standard combination therapy with beta-lactam antibiotics for severe community-acquired pneumonia^8^, owing to its ability to suppress inflammatory cytokines during these critical cases. Viral respiratory diseases, such as influenza, are not an exception in the mechanism of exacerbation, and combination therapy with clarithromycin and antiviral agents showed clinical efficacy in influenza A infection ^9, 10^. Regarding the use of macrolides for COVID-19, a combination therapy of azithromycin and hydroxychloroquine was reported as effective for viral elimination ^11^, although another case series reported no benefit in severe COVID-19 patients^12^. Nevertheless, these reports did not investigate the efficacy of macrolide monotherapy, and no study has used macrolide to target mild COVID-19. Since azithromycin has a risk for circulatory side effects, this remains a concern for its use in treating COVID-19 patients who often have cardiovascular comorbidities. Clarithromycin is a good candidate for preventing the exacerbation of COVID-19 by suppressing inflammatory cytokines and may be safely used in patients with COVID-19. This trial is planned to estimate the efficacy of clarithromycin in patients with mild COVID-19 pneumonia who do not require oxygen administration.

## Methods and Analysis

### Study design and setting

“The <u>CAM</u> (Clarithromycin) <u>E</u>ffectivity for <u>COVID</u>-19 pneumonia which does not require oxygen administration; multicenter, randomized-controlled, open-label, 3-armed parallel group comparison, exploratory trial” (CAME COVID study) was initiated in May 2021, after the approval by the Clinical Research Review Board in Nagasaki University in March, 2021 and the registration/publication at the Japan Registry of Clinical Trials (jRCT) (registration number: jRCTs071210011) in April, 2021. This study was planned to end in July 2022. Subjects will be enrolled from May, 2021 and enrolment will end in February 2022. As shown in Figure 1, subjects who are eligible for this study will be asked to participate in this study, and informed consent will be obtained prior to the registration/randomization. After written consent is obtained from the eligible subjects, they will be enrolled and randomized into Group A (administration of clarithromycin 800 mg/day), Group B (administration of clarithromycin 400 mg/day), or Group C (standard treatment without clarithromycin). The rationale for the doses of clarithromycin is as follows: 400 mg/day and 800 mg/day of clarithromycin have been approved for bacterial respiratory infectious diseases and nontuberculous mycobacterial infection, respectively, in Japan, and the safety of these doses of clarithromycin has been confirmed.

**Figure 1.**
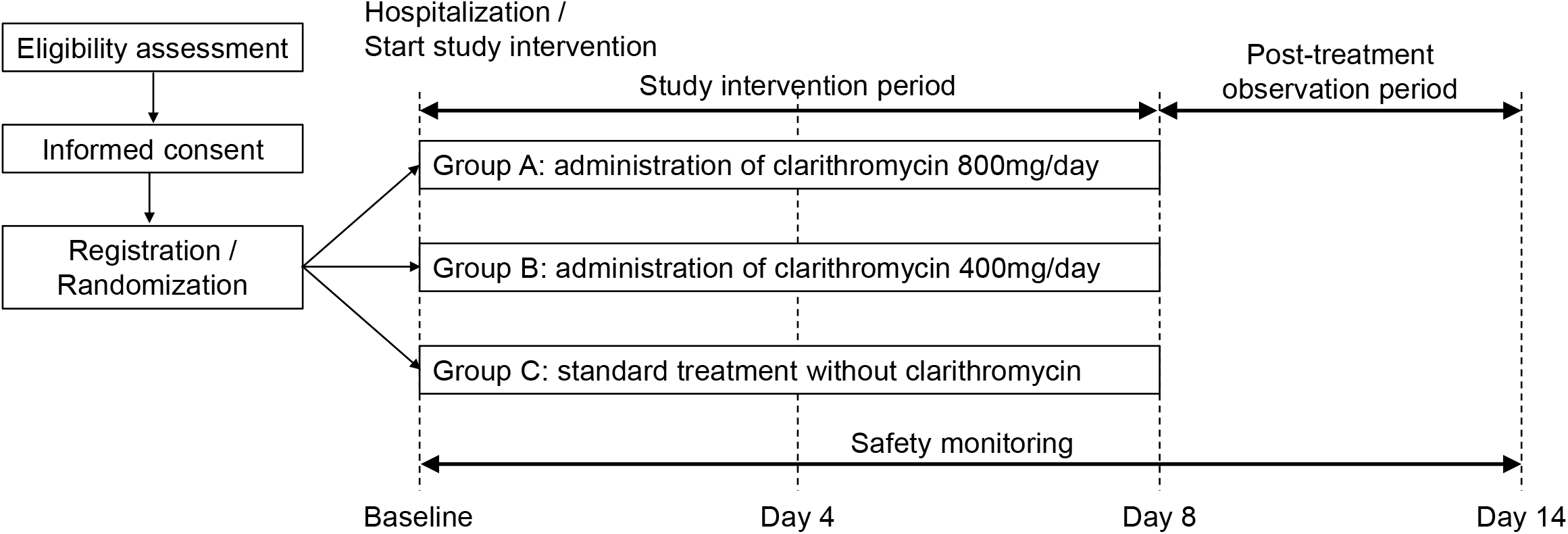
Study design and flow of recruitment, randomization, study intervention and observation.

### Sample size calculation

Since no clinical trial has evaluated the effect of clarithromycin on COVID-19 pneumonia, no reference data are available for the statistical sample size calculation in this study. Therefore, this study was planned as an exploratory trial, and the target number of enrolled subjects was defined as 60 (20 subjects × 3 groups), based on the possible number of subjects who could give their consent during the planned enrolment period in this study at the participating medical institutions.

### Eligibility criteria

Patients with COVID-19 pneumonia who do not require oxygen administration will be included in this study. The detailed inclusion criteria are as follows: 1) patients in whom SARS-CoV-2 is detected by PCR tests or loop-mediated isothermal amplification method within 3 days before the informed consent; 2) patients with pneumonia by routine chest radiography or chest computed tomography (CT); 3) Japanese who are aged 20 years or older; and 4) patients who give their written consent form to participate in the study. The exclusion criteria are as follows: 1) patients who had symptoms of 8 days or longer; 2) patients who were treated with macrolide antimicrobial agents; 3) patients who were treated with steroids (except inhalants) or immunosuppressive agents; 4) patients who are diagnosed with influenza viral infection; 5) patients whose percutaneous arterial oxygen saturation is 94% or less under room air condition; 6) patients with hepatic dysfunction (aspartate aminotransferase or alanine aminotransferase is more than five times the upper limit of normal in each medical institution or Child-Pugh B or C); 7) patients with renal dysfunction. (creatinine is more than twice the upper limit of normal in each medical institution, and estimated glomerular filtration rate is less than 30 ml/min/1.73 m^2^); 8) patients whose peripheral blood neutrophils are less than 1,000/µL; 9) patients who have a history of hypersensitivity to macrolide antimicrobial agents; 10) patients who are pregnant or breastfeeding; 11) patients who have a history of vaccination against COVID-19; and 12) patients with other conditions that the investigator thinks may render them inappropriate to participate in the study.

### Recruitment and consent

The informed consent document will be offered to the candidates who meet all of the inclusion criteria and who do not fail any of the exclusion criteria to provide a comprehensive explanation of this study. Written consent will be obtained. After obtaining consent, the participants are enrolled in this study.

### Random allocation

After obtaining informed consent, eligible subjects will be randomly assigned in a 1:1:1 ratio to Group A (administration of clarithromycin 800 mg/day), Group B (administration of clarithromycin 400 mg/day), or Group C (standard treatment without clarithromycin). The randomization sequence was generated using a computer-based dynamic allocation method with a minimization procedure to balance the allocation factors (age ≥ 20 years and less than 40 years, ≥ 40 years and less than 60 years, or ≥ 60 years) and sex.

### Study intervention and observation

All enrolled subjects in this study will be hospitalized for 7 days ^13^ for a comprehensive follow-up during the intervention. Subjects assigned to Group A will start to take 400 mg of clarithromycin orally twice daily (after breakfast and supper) after hospitalization. Subjects assigned to Group B will start to take 200 mg of clarithromycin orally twice daily (after breakfast and supper) after hospitalization. Subjects assigned to Group C will start to take the standard care for COVID-19 pneumonia in each medical institution without the administration of clarithromycin after hospitalization. In groups A and B, clarithromycin will be administered 14 times (twice daily for 7 days). If the administration starts on the morning of day 1 (the day of hospitalization), clarithromycin is administered until the evening of day 7. If the administration started on the evening of day 1, clarithromycin is administered until the morning of day 8. The study subjects will be observed on days 1, 2, 3, 4, 5, 6, 7, 8, and 14. The day of hospitalization is considered to be the day when observation starts (reference date). During the study intervention period (days 1 to 7 or 8), the subjects will not be allowed to use macrolide antimicrobial agents, steroids, or immunosuppressive agents. After completion of the study intervention (day 8), the study subjects will be discharged. After day 8 (post-treatment observation period), all study subjects will receive standard care for COVID-19 pneumonia in each medical institution without the administration of clarithromycin.

Table 1 shows the schedule of assessments performed at each observation point, including the mandatory and optional assessments. 2) Inspection of subjects’ characteristics (height, weight, body mass index, onset of COVID-19, detection date of SARS-CoV-2, date of hospitalization, anamnesis and comorbidity); 4) vital signs (body temperature, systolic blood pressure, diastolic blood pressure, pulse, percutaneous oxygen saturation, and frequency of breath); 7) hematology tests (red blood cell, hemoglobin, hematocrit, white blood cell, neutrophil, lymphocyte, eosinophil, monocyte, basophil, and platelet); 8) general blood biochemical tests (total bilirubin, aspartate aminotransferase, alanine aminotransferase, alkaline phosphatase, gamma-guanosine triphosphate, total cholesterol, total protein, albumin, blood urea nitrogen, creatinine-estimated glomerular filtration rate based on creatinine, lactate dehydrogenase, creatine phosphokinase, brain natriuretic peptide, troponin T, C-reactive protein, procalcitonin, ferritin, Na, pH, and hemoglobin A1c); 9) blood coagulation tests (prothrombin time, activated partial thromboplastin time, and d-dimer); 10) chest radiography and computerized tomography; and 14) inspection of medications of other pharmaceutical agents are conducted by general inspection and interview. 3) The severity of COVID-19 pneumonia is measured by the severity classification according to the COVID-19 infectious disease treatment guidelines by the Ministry of Health, Labour and Welfare in Japan^2^, pneumonia severity index ^14^, and A-DROP defined in the guidelines for the management of community-acquired pneumonia in adults released from the Japanese Respiratory Society^15^; 5) the quantity of oxygen administered will be recorded daily during hospitalization; 6) PCR tests for SARS-CoV-2 will be conducted using nasopharyngeal swabs; 11) nasal drip tests will be conducted for interleukin (IL)-1beta, IL-6, IL-8, IL-10, IL-17, tumor necrosis factor-alpha, interferon-gamma, beta-defensin, granulocyte-macrophage colony-stimulating factor, and immunoglobulin A; 12) special blood tests will be conducted for cytokines, chemokines, IL-33, immunoglobulin M, immunoglobulin G, and immunoglobulin A; 13) medication adherence of the study agent; 15) meal intake; and 16) subjective symptoms measured by the Severity Score^16^ are daily answered on the study subjects’ diary by the study subjects themselves.

**Table 1.**
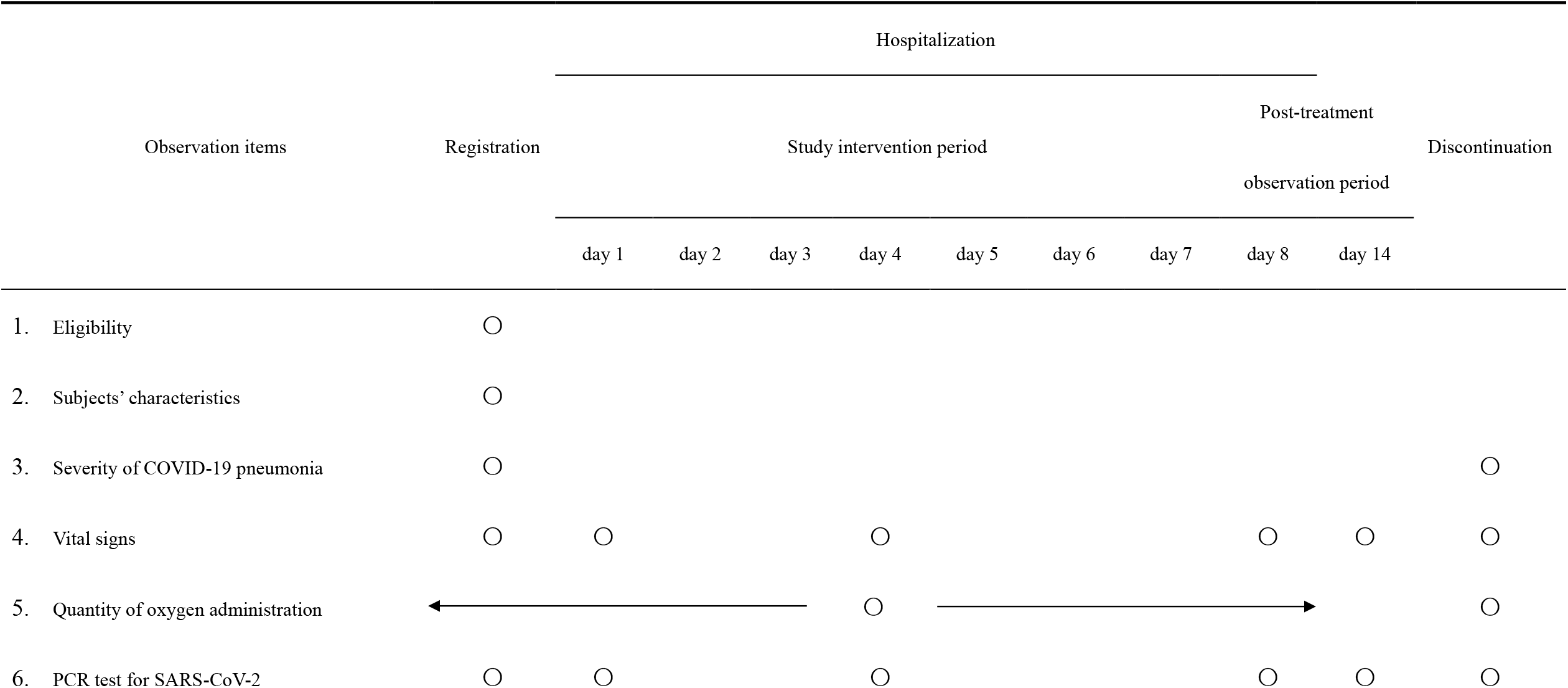
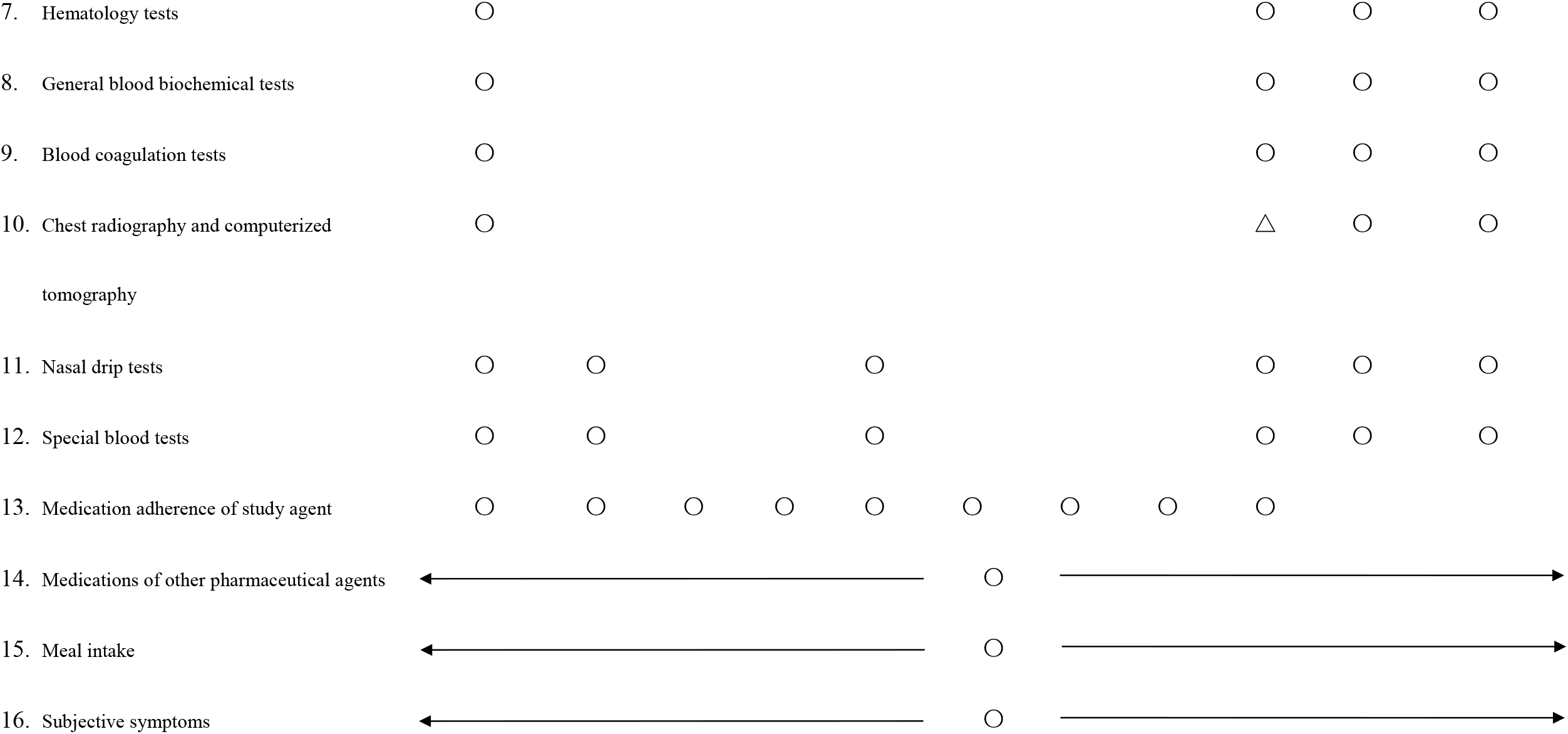
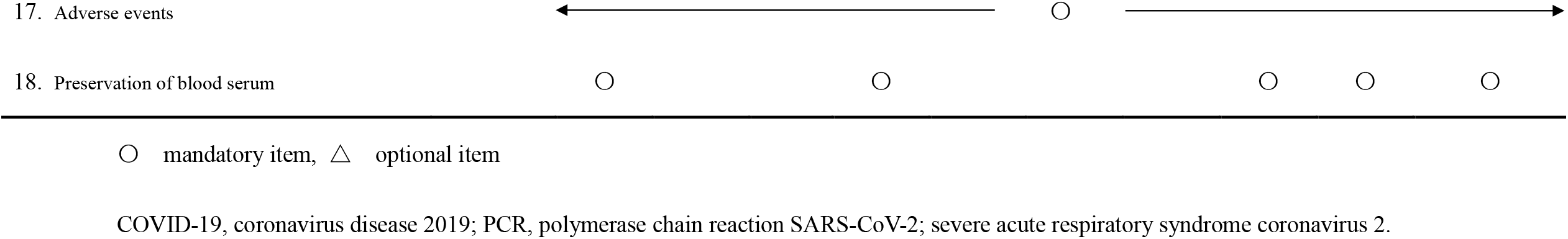
Schedule of assessments performed at each observation point

### Outcomes

The primary endpoint of this study will be the number of days required to improve clinical symptoms, measured by the Severity Score^16^, by 50% or more from baseline. Secondary endpoints will be as follows: 1) days to recover the body temperature to below 37°C after registration; 2) change in inflammatory cytokines in serum and nasal discharge from baseline; 3) proportion of subjects in whom all the clinical symptoms measured by the Severity Score completely disappeared; 4) proportion of subjects in whom each clinical symptom measured by the Severity Score is improved by 50% or more from baseline; 5) reduction rate of the SARS-CoV-2 from baseline; 6) change in serum immunoglobulin G, immunoglobulin M, and immunoglobulin A antibodies from baseline; 7) recovery rate of peripheral blood lymphocytes; 8) proportion of subjects with oxygen administration; 9) change in other general blood biomarkers from baseline; and 10) change in pneumonia image by chest radiography or chest CT. The Severity Score ^16^ is measured by asking the effect of five pneumonia-related symptoms (cough, shortness of breath, fatigue, headaches, and anosmia) on the subjects’ daily life on a 4-point Likert scale (1 = not affected, 2 = little affected, 3 = affected, and 4 = severely affected).

### Data collection, data management, and monitoring

A case report form will be used for the data collection. A central registration number will be used to identify the participants for anonymization. Data collection and management will be carried out by third-party entities to avoid bias. Data management will be performed by Soiken Inc., Data Management Group (Data Center). To manage and ensure quality, the study is monitored by Soiken Inc., Monitoring Group.

### Safety evaluation

During this study, the investigators constantly monitored any adverse events (AEs) through regular medical checkups. All related AEs, not only side effects to the study agents but also any abnormal clinical laboratory test values and any untoward medical occurrence, are reported and documented. If the AEs meet the following criteria, the events are referred to as serious adverse events (SAEs), based on the ICH E2A, ICH E2D, and the “Ethical Guidelines for Medical and Health Research Involving Human Subjects” in Japan^17^: 1) AEs that result in death; 2) AEs that are life-threatening; 3) AEs that require hospitalization or prolongation of existing hospitalization; 4) AEs that result in persistent or significant disability or incapability; 5) other AEs that are medically important or critical; 6) AEs that are equivalently severe to criteria 1) and 5); and 7) AEs that are congenital normality or birth defects. AEs were followed up until normalization or recovery to a level not considered to be an adverse event, or until four weeks after the end of the pre-observation period or study discontinuation. The first hospitalization after the registration to day 8 and prolongation of the first hospitalization after day 8 without worsening of pneumonia-related symptoms were not considered as AEs/SAEs in this study, since this study enrolled COVID-19 patients with pneumonia images who should be treated by hospitalization until the complete disappearance of pneumonia-related symptoms, according to the COVID-19 infectious disease treatment guidelines by the Ministry of Health, Labour and Welfare in Japan^2^.

### Statistical analysis

All tests will be two-sided, and a *p* value of < 0.05 will be considered statistically significant. A statistical analysis plan will be developed prior to the database lock. All statistical analyses will be conducted by independent biostatisticians.

Three analysis sets were defined in this study; the full analysis set (FAS) included all subjects who were registered in this study and assigned to one of the intervention groups. However, subjects with a severe protocol violation, such as registration without consent or registration out of the enrollment period, will be excluded from the FAS. The per-protocol set (PPS) excludes subjects with a protocol violation from the FAS, such as violation of eligibility criteria, use of prohibited or restricted concomitant drugs, or poor adherence to the study agent (less than 75% or more than 120%). The safety analysis set included all subjects who were registered in this study and received at least one dose of the study agent or control treatment.

Subject characteristics at baseline will be presented as frequencies and proportions for categorical data, and summary statistics (number of subjects, mean, standard deviation, minimum, median, and maximum) for continuous data. Subject characteristics will then be compared using the chi-square test or Fisher’s exact test for categorical endpoints, and analysis of variance or Kruskal-Wallis test for continuous variables.

The primary endpoint of this study is the number of days required to improve clinical symptoms measured by the Severity Score^16^ by 50% or more from baseline. The primary endpoint will be analyzed using the FAS and PPS. The cumulative incidence curves for the days required to improve clinical symptoms measured by the severity score will be drawn, and a log-rank test with a closed testing procedure will be conducted for comparison among the three groups. In addition, hazard ratios among the groups will be estimated using the Cox proportional hazard model, and a 95% confidence interval will be calculated. The hazard ratio adjusted by allocation factors (age and sex) as covariates will also be estimated.

For the secondary endpoints, summary statistics (number of subjects, mean, standard deviation, minimum, median, and maximum) for measurements, changes from baseline, and percent changes from baseline will be calculated for continuous data. Frequencies and proportions will be calculated for the categorical data. Two-sample *t*-test or Wilcoxon signed-rank test for intergroup comparisons of the continuous data, one-sample *t*-test or Wilcoxon signed-rank test for intragroup comparisons of the continuous data, and the chi-square test or Fisher’s exact test for intergroup comparisons of the categorical data will be performed. Multiplicity will not be adjusted for the secondary endpoints.

For the safety endpoints, summary statistics for the frequency of adverse events will be calculated, and Fisher’s exact tests will be performed for intergroup comparisons. Multiplicity will not be adjusted for the safety endpoints.

### Strength and limitations of the study

This is the first randomized controlled trial to evaluate the efficacy of clarithromycin against COVID-19 pneumonia. Although several *in vitro* or bioinformatics analyses have reported the positive effect of macrolide antimicrobial agents on coronaviruses, no clear clinical evidence has confirmed the efficacy of macrolides, including clarithromycin, on coronaviruses, especially SARS-CoV-2^18^. There is no treatment strategy established to prevent exacerbation in patients with mild COVID-19 who do not require oxygen administration. Development of a proper treatment strategy for patients with mild COVID-19 is important, not only for the medical aspects, but also in social and economic aspects, such as, for example, to reduce the hospital bed occupancy by exacerbated COVID-19 patients or to suppress the treatment costs for patients with severe COVID-19. Clarithromycin may also contribute to the prevention of secondary infections due to its antiviral effects. Therefore, this study aimed to evaluate the efficacy of clarithromycin in patients with mild COVID-19 pneumonia who did not require oxygen administration.

The results of this study may contribute to the development of new treatment strategies for COVID-19 pneumonia.

Our study has several limitations. First, it is exploratory in nature. Because of the lack of previous clinical evidence that evaluated the effect of clarithromycin on COVID-19 pneumonia, the target number of enrolled subjects was defined based on the feasible number of subjects who could give their consent during the planned enrolment period in this study at the participating medical institutions. Second, the target sample size was relatively small. Therefore, if the improvement of pneumonia-related symptoms by clarithromycin is relatively smaller than expected, power will be insufficient to detect significant intergroup differences. Third, this was an open-label trial. Thus, bias for the subjects and investigators/physicians cannot be completely avoided. Fourth, this study was conducted only in medical institutions in Japan and enrolled only Japanese patients. These constraints may limit the generalizability of this study. Further international clinical trials on a larger scale are required in the future.

### Patient and public involvement

The Patients and the public are not involved in the study, including planning, execution, analysis, and evaluation.

### Ethics and dissemination

This study and its protocol were approved by the Clinical Research Review Board of Nagasaki University (approval no. CRB20-027) in accordance with the Clinical Trials Act of Japan. The study was conducted in accordance with the Declaration of Helsinki, the Clinical Trials Act, and other current legal regulations in Japan. Written informed consent was obtained from all participants after a full explanation of the study. The results of this study will be disseminated at medical conferences and through journal publications.

## Data Availability

No data is available public after the completion of this study, because the study protocol does not include the statement to share the data public, and the ethics committee did not approve the data sharing.

## Acknowledgments

The authors thank all the clinical staff for their assistance in the execution of the study and Soiken Inc. for their technical assistance in the launch and execution of the study.

The study is funded by Taisho Pharmaceutical Co., Ltd.

## Author contributions

Kazuko Yamamoto and Naoki Hosogaya contributed to the conception and design of the study, drafted the protocol, and supervised the revision. Noriho Sakamoto, Haruo Yoshida, Kenji Ota, Hiroshi Ishii, Kazuhiro Yatera, Koichi Izumikawa, and Katsunori Yanagihara provided intellectual input to improve the study design and revise the protocol. Hiroshi Mukae supervised the conception and design of this study. All authors read and approved the final manuscript.

